# Development and External Validation of a Plasma p-tau217–Based Model for MCI-to-Dementia Progression

**DOI:** 10.64898/2026.09.15.26363045

**Authors:** Meixi Du, Ishaanee Roy, Berne Chu, Shujin Tian, Alzheimer’s Disease Neuroimaging Initiative

## Abstract

—Clinical trajectories after a diagnosis of mild cognitive impairment (MCI) vary widely, so prognostic models must account for both the timing of subsequent diagnoses and unequal follow-up. Using the Alzheimer’s Disease Neuroimaging Initiative (ADNI), we developed a Cox proportional-hazards model for time from an MCI-aligned plasma index visit to a subsequent dementia diagnosis, using age at study entry and log-transformed plasma p-tau217. Coefficients and baseline survival were estimated in 401 participants with 85 events and then frozen and applied without refitting to an independent cohort from the National Alzheimer’s Coordinating Center (NACC). NACC p-tau217 measurements were reported on the Quanterix HD-X platform and were mapped to the ADNI development-assay scale using a prespecified transformation derived from paired ADNI measurements. The two-predictor model reached a pooled out-of-fold C-index of 0.775 in ADNI (apparent C-index, 0.778) and 0.652 (95% CI, 0.526–0.765) in 104 NACC participants with 25 incident dementia diagnoses. At two years, observed risk was 0.214 compared with a mean predicted risk of 0.089, an observed-to-expected ratio of 2.40. Neither adding APOE *ε*4 count, sex, or education nor expanding the model with GFAP, NfL, and the A*β*42/A*β*40 ratio materially improved discrimination in ADNI; APOE *ε*4 also provided no detectable incremental gain in NACC. The model showed moderate discrimination in NACC, although the wide confidence interval includes values near chance and the middle and higher risk tertiles did not clearly separate; C-indices were similar in the bridged primary and direct-scale sensitivity analyses, and mean two-year predicted risk was lower than observed only in the bridged analysis.

## I. Introduction

Clinical trajectories after a diagnosis of mild cognitive impairment (MCI) vary substantially: some individuals receive a dementia diagnosis within a short interval, whereas others remain without such a diagnosis for years [1]. Prognostic models should therefore account for both the timing of subsequent diagnoses and unequal follow-up, motivating a time-to-event approach.

Blood-based biomarkers offer a more accessible approach to risk assessment than cerebrospinal fluid or imaging measures [2], [3]. Plasma phosphorylated tau 217 (p-tau217) is of particular interest because it reflects Alzheimer disease pathology [3]–[5] and has been associated longitudinally with progression to dementia or cognitive impairment [6], [7]. Prior prognostic studies in MCI have evaluated combinations of multiple plasma biomarkers [8]. However, whether an age–p-tau217 model retains its discrimination and calibration when transferred to an independent cohort remains uncertain.

We therefore aimed to develop and internally validate in the Alzheimer’s Disease Neuroimaging Initiative (ADNI) a Cox model using age and plasma p-tau217 to estimate time to a subsequent dementia diagnosis, and then to externally validate the frozen model in an independent cohort from the National Alzheimer’s Coordinating Center (NACC). In supporting analyses, we also assessed the incremental value of apolipoprotein E (APOE) *ε*4 count, sex, education, and additional plasma biomarkers.

## II. Materials and Methods

### A. Cohorts and Outcome

Data used in the preparation of this article were obtained from the Alzheimer’s Disease Neuroimaging Initiative (ADNI) database (adni.loni.usc.edu). The ADNI was launched in 2003 as a public-private partnership, led by Principal Investigator Michael W. Weiner, MD. The primary goal of ADNI has been to test whether serial magnetic resonance imaging (MRI), positron emission tomography (PET), other biological markers, and clinical and neuropsychological assessment can be combined to measure the progression of mild cognitive impairment (MCI) and early Alzheimer’s disease (AD).

This study developed a prognostic model in ADNI [9] and evaluated the frozen model externally in NACC.

For the primary development analysis, four participant-level ADNI sources provided longitudinal diagnoses, plasma biomarkers, APOE genotype, and age at study entry; the demographic extension additionally used the ADNI demographic case report form.

ADNI participants were anchored at the earliest plasma draw whose nearest diagnosis within *±*90 days was MCI. Participants without subsequent diagnostic follow-up or complete candidate-model predictors were excluded, yielding the common development cohort (Fig. 1).

**Fig. 1.**
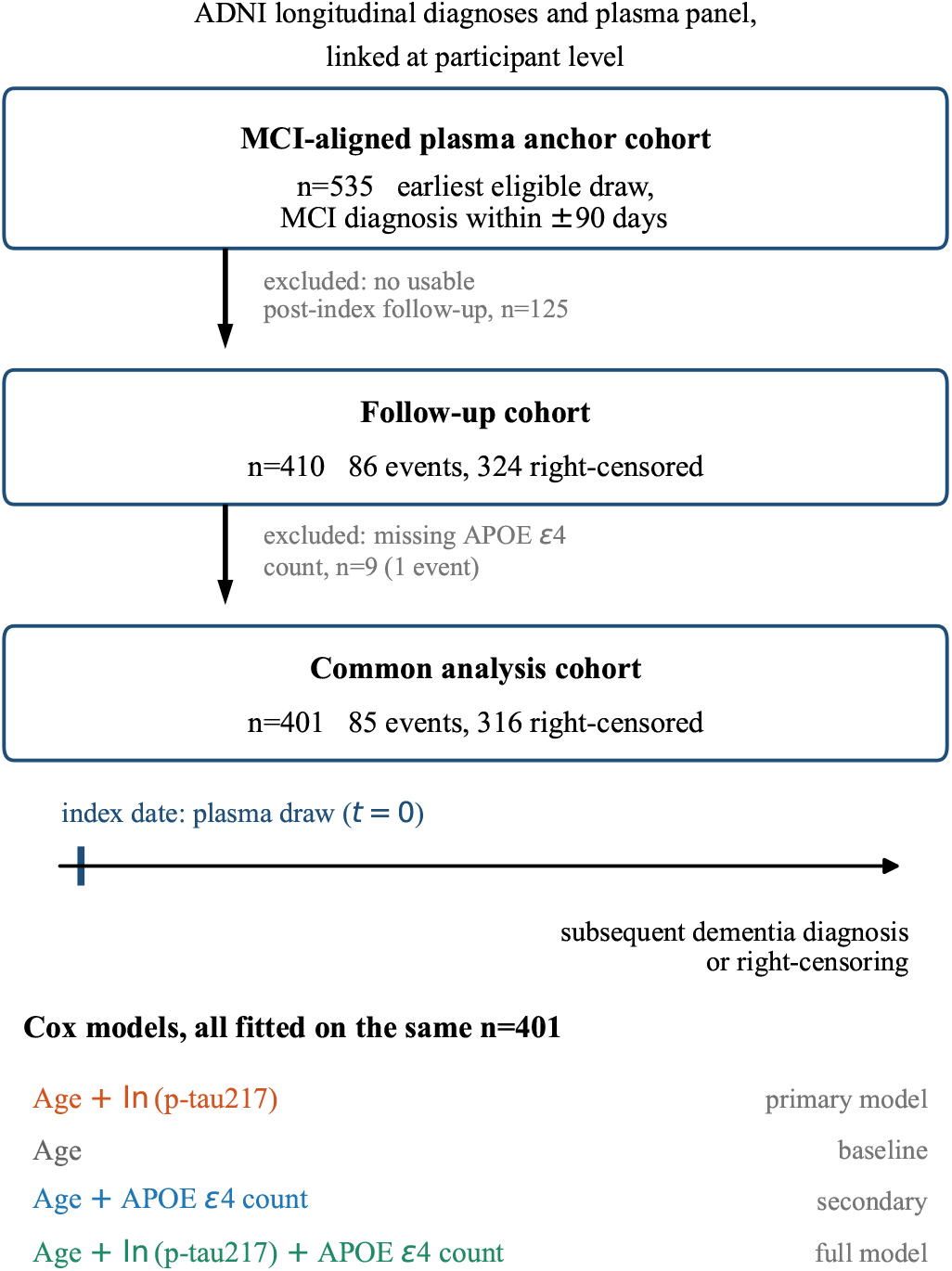
ADNI cohort derivation and study design. All four Cox models were fitted and evaluated on the same 401-participant common analysis cohort. These were the original development comparisons; additional demographic/genetic and biomarker extensions were evaluated separately as supporting analyses.

External validation linked National Alzheimer’s Coordinating Center (NACC) Uniform Data Set visits to plasma biomarker measurements. The longitudinal clinical data came from the NACC Uniform Data Set. The plasma p-tau217 values were not obtained from NCRAD; they were obtained from the February 2026 ADSP Phenotype Harmonization Consortium (ADSP-PHC) harmonized data release (ng00067.v20) through an approved NACC Quick Access File data request. NACC was not involved in the harmonization of these biomarker data. Participants were anchored at the biomarker-linked MCI visit and required no recorded dementia on or before it, complete age and p-tau217, and subsequent followup. Because specimen dates were unavailable, the linked visit date served as the index date.

In both cohorts, the event was the first recorded dementia diagnosis after the index date; participants without an event were censored at their last eligible nondementia visit. Deaths were treated as censoring at the last recorded nondementia visit rather than modelled as competing events. No minimum follow-up was imposed. The endpoint was dementia of any cause because too few events had etiologic diagnoses to support an Alzheimer-specific outcome.

### B. Predictors and Frozen Model

ADNI plasma p-tau217 was measured on the Fujirebio Lumipulse platform in pg/mL and entered all models as the natural logarithm of the raw value. APOE *ε*4 was coded as an allele count of 0, 1 or 2. Age was age at study entry in both cohorts.

The primary model contained age at study entry and log-transformed plasma p-tau217. Its coefficients and baseline survival were estimated in ADNI and frozen before external validation in NACC.

Supporting ADNI analyses considered sex, education, pre-index MMSE and FAQ, log-transformed GFAP and NfL, and the A*β*42/A*β*40 ratio. Sex and education came from the ADNI demographic case report form with male as the reference category, and were missing for 10 of 401 participants (2.5%), handled by fold-specific imputation with a missingness indicator. All other missing-data processing was confined to training folds. None of these variables entered the primary frozen model or determined primary NACC eligibility.

### C. Assay Harmonisation

NACC p-tau217 measurements were reported on the Quan-terix HD-X platform and were mapped to the Fujirebio-equivalent scale required by the frozen model using a pre-specified bridge derived from paired ADNI measurements on both platforms,

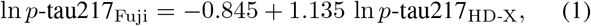

estimated by least squares on the logarithmic scale, both sides in pg/mL. It was fitted to 1,175 paired measurements from 393 ADNI participants (up to three draws each) spanning 0.032–6.296 pg/mL on the Quanterix side and 0.030–3.170 pg/mL on the Fujirebio side; under participant-level cross-validation *R*^2^ was 0.725 and the residual standard deviation 0.485 on the log scale. All NACC values fell within 0.032–6.296 pg/mL, the range spanned by the paired measurements, so no extrapolation was required.

### D. Statistical Analysis

All models were Cox proportional-hazards regressions [11]. Four candidate models were fitted on the identical 401-participant development cohort (Fig. 1); the primary comparison was the age-only model against the age-plus-p-tau217 model, and the APOE-containing models supported secondary comparisons. The hazard ratio associated with a twofold increase in p-tau217 was calculated as 2^*β*^, where *β* is the coefficient for log-transformed p-tau217. Discrimination was assessed using five-fold cross-validation repeated five times, with all preprocessing confined to the training folds, and summarised by Harrell’s concordance index (C-index) [12], two-year time-dependent area under the receiver operating characteristic curve and Brier score, with 95% confidence intervals from a paired participant-level bootstrap of 2,000 replicates; apparent performance is reported separately from out-of-fold performance. Nested models were compared by likelihood-ratio test and the Akaike information criterion (AIC), and proportional hazards from scaled Schoenfeld residuals [13]. Out-of-fold calibration at 2 and 4 years used jackknife pseudo-observations with a complementary log-log link [14], [15].

The age-plus-p-tau217 model was selected during model development and was not prespecified. Two supporting analyses used the additional predictors defined in Section II-B within the same cross-validation and bootstrap framework. In a pre-specified post-development extension, the demographic terms were added individually and jointly to the retained model. In a separate supporting clinical benchmark, a model containing age, APOE *ε*4 count, the pre-index cognitive and functional measures and log-transformed p-tau217 was compared with an expanded model adding the three remaining plasma biomarkers, in the same 401 participants using fold-specific imputation and no follow-up information in preprocessing. Further sensitivity analyses varied landmarking, anchor alignment, outcome definition and the functional form of p-tau217, and assessed the influence of individual observations.

In NACC the frozen model was applied without refitting or recalibration. We report Harrell’s C-index; two-year time-dependent area under the receiver operating characteristic curve and inverse probability of censoring weighted Brier score; Kaplan–Meier observed two-year risk; and the observed-to-expected ratio, observed divided by mean predicted risk. A calibration slope from the frozen linear predictor is reported as a diagnostic only. Uncertainty came from participant-level bootstrap resampling, with the external C-index interval additionally incorporating bridge uncertainty by re-estimating (1) within each resample. Three further analyses left the cohort, the outcome and the evaluation procedure unchanged, and none refitted or recalibrated the model: a measurement-scale sensitivity analysis treating NACC values as directly equivalent to Fujirebio concentrations; a secondary comparison, on the primary bridged scale, of the frozen model against one additionally containing APOE *ε*4 allele count in the 103 participants with complete counts; and a post hoc sensitivity analysis replacing study-entry age with age at the index date, reconstructed from study-entry age and elapsed time to the plasma index in ADNI and taken from the anchor visit in NACC. The two paired comparisons each used 2,000 participant-level bootstrap samples. Reporting follows guidance for prediction-model studies [10].

## III. Results

### A. Development in ADNI

The common analysis cohort comprised 401 participants with 85 subsequent dementia diagnoses and 316 right-censored observations (Fig. 1). Table I reports the characteristics of this cohort alongside those of the external cohort. In the age-plus-p-tau217 model, plasma p-tau217 carried a hazard ratio of 2.14 per twofold increase (95% CI 1.79–2.56, *p <* 0.001). Adding ln p-tau217 to the age-only baseline improved model fit substantially (likelihood-ratio *χ*^2^ = 68.6 on 1 degree of freedom, *p <* 0.001; AIC 806.3 to 739.7).

**TABLE I.** Baseline characteristics of the development and external cohorts.

|  | ADNI<br>(n=401) | NACC<br>(n=104) |
| --- | --- | --- |
| Age at study entry, y | 72.7 [67.6–77.5] | 71.0 [65.0–77.3] |
| Age at index visit, y | 74.8 [69.2–80.0] | 73.5 [67.0–80.0] |
| Female sex, n (%) | 162 (41.4%) | 51 (49.0%) |
| Education, y | 16.0 [14.0–18.0] | 16.0 [14.0–18.0] |
| APOE ε4 alleles, n (%) | | |
| 0 | 215 (53.6%) | 58 (56.3%) |
| 1 | 144 (35.9%) | 34 (33.0%) |
| 2 | 42 (10.5%) | 11 (10.7%) |
| MMSE (ADNI, pre-index) | 28.0 [26.0–29.0] | – |
| MoCA (NACC, index visit) | 23.0 [21.0–25.0] | 24.0 [21.0–25.0] |
| FAQ total | 2.0 [0.0–5.0] | 1.0 [0.0–5.0] |
| p-tau217, pg/mL, native | 0.215 [0.119–0.479] | 0.390 [0.230–0.753] |
| bridged to Fujirebio scale | – | 0.148 [0.081–0.311] |
| Follow-up, d | 425 [285–1121] | 661 [421–765] |
| Incident dementia, n (%) | 85 (21.2%) | 25 (24.0%) |
| Deaths during follow-up, n (%) | – | 3 (2.9%) |

Continuous variables are median [interquartile range]; categorical variables are n (%), over participants for whom the variable was observed. Missing: Sex (ADNI 10); Education (ADNI 10); APOE $\epsilon$ 4 count (NACC 1); MMSE (ADNI 64); MoCA (ADNI 101, NACC 3); FAQ total (ADNI 81, NACC 38). p-tau217 is on each cohort's native platform (ADNI, Fujirebio Lumipulse; NACC, Quanterix HD-X) and, for NACC, also after (1). MMSE and MoCA are the latest ADNI pre-index values and the NACC index-visit values; no NACC index visit records an MMSE, each having used a form version administering the MoCA. The NACC FAQ total sums the ten Uniform Data Set B7 items. Deaths are counted from the index date onward.

The frozen fit is reported in full so that a predicted risk can be recomputed from this paper alone. The coefficient for age at study entry was 0.0154 per year (standard error, *−* 0.0170; 95% CI, 0.0179 to 0.0487; hazard ratio, 1.015; 95% CI, 0.982–1.050) and for ln p-tau217 it was 1.097 (standard error, 0.1309; 95% CI, 0.840 to 1.354; hazard ratio, 2.995; 95% CI, 2.317–3.871). Predicted risk by time *t* is

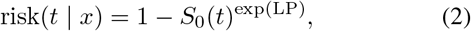

with the linear predictor centred on the development-cohort means,

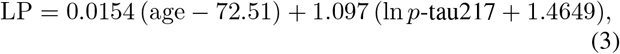

where age is in years and p-tau217 is in pg/mL on the Fujirebio scale, and with frozen baseline survival *S*_0_(*t*) = 0.9669, 0.9080 and 0.6660 at one, two and four years. Together with (1), these values fully specify the model as it was applied in NACC.

The age-plus-p-tau217 model reached a pooled out-of-fold C-index of 0.775 against 0.557 for the age-only model, a paired difference of +0.218 (95% CI +0.136 to +0.295), and an apparent C-index of 0.778 (Fig. 2(a)). Adding APOE *ε*4 allele count changed out-of-fold discrimination by *−*0.001 (95% CI *−*0.005 to +0.003), did not improve fit (*p* = 0.77) and raised AIC to 741.6; the interval is narrow and centred near zero, but does not establish equivalence.

**Fig. 2.**
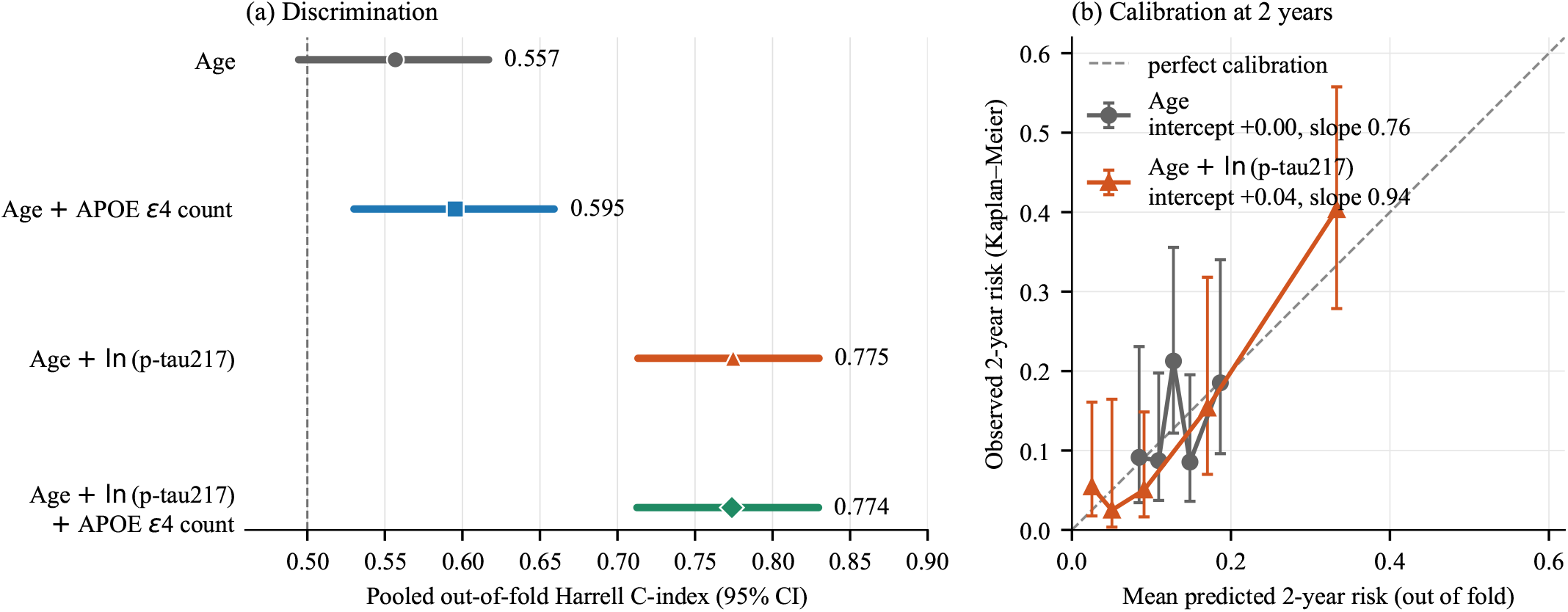
Out-of-fold performance of the original development models in ADNI (*n*=401; 85 events). (a) Pooled Harrell C-index with paired-bootstrap 95% confidence intervals; the dashed line marks chance. (b) Two-year calibration for the age-only and age-plus-p-tau217 models, showing quintile-level mean predicted and Kaplan–Meier observed risk with 95% confidence intervals.

In the prespecified post-development extension, adding sex and education did not improve out-of-fold discrimination beyond age and log-transformed p-tau217 (ΔC = *−*0.0005; 95% CI, *−*0.0119 to 0.0113); neither term was individually associated with progression (sex hazard ratio, 0.85; 95% CI, 0.52–1.39; *p* = 0.52; education hazard ratio per year, 1.05; 95% CI, 0.97–1.15; *p* = 0.22). The interval excludes a material improvement larger than approximately 0.011 in C-index, although very small effects remain compatible with the data. By contrast, adding p-tau217 to a demographic baseline containing age, sex and education improved discrimination by 0.235 (95% CI, 0.155 to 0.310).

In the separate supporting clinical benchmark, expanding the p-tau217–containing model with GFAP, NfL and A*β*42/A*β*40 did not improve out-of-fold discrimination (C-index, 0.791 versus 0.789; paired ΔC = *−*0.002; 95% CI, *−* 0.016 to +0.013; *n*=401, 85 events).

Out-of-fold calibration at 2 years gave an intercept of +0.04 and a slope of 0.94 (95% CI 0.50 to 1.98) with an observed-to-expected ratio of 1.00 (Fig. 2(b)); the intervals are wide, indicating no detectable miscalibration rather than precise calibration. At 4 years the slope was 1.34 and the ratio 0.99. There was no evidence of violation of the proportional-hazards assumption for either term (*p ≥* 0.38). The discrimination advantage over the age-only model persisted across every supporting analysis: landmarking, all nested anchor-alignment restrictions and the confirmed-conversion endpoint each gave paired differences whose intervals excluded zero, and a restricted cubic spline did not improve on the linear term.

### B. External Validation in NACC

Of 692 participants with a measured p-tau217 value in the February 2026 ADSP-PHC harmonized plasma release (ng00067.v20), 633 linked to a Uniform Data Set clinical visit (59 excluded for having neither a specimen-date nor a visit-number match), 126 were at a visit with MCI and no dementia recorded on or before it (507 excluded), all 126 had complete age and p-tau217 (none excluded), 108 had at least one subsequent follow-up visit (18 excluded), and 104 linked by an exact visit number rather than an approximate match (4 excluded).

The resulting external cohort comprised 104 participants with MCI, of whom 25 received an incident dementia diagnosis; median follow-up was 661 days (interquartile range, 421–765). On the bridged scale the NACC p-tau217 distribution sat below the development distribution (median, 0.148 versus 0.215 pg/mL; Table I), yet a slightly larger share of participants received a dementia diagnosis (25 of 104 versus 85 of 401) — lower predicted risk alongside at least as much observed progression, the direction of the calibration gap reported below. Eligibility required only the two predictors the model uses, so APOE genotype was not required for primary-cohort eligibility. Applying the frozen two-predictor model gave a C-index of 0.652 (95% CI, 0.526–0.765), an interval that includes uncertainty in the assay bridge. Cumulative dementia incidence was ordered as expected between the lower-risk tertile and the two higher tertiles, but the middle and higher tertiles overlapped throughout and crossed after approximately 1.5 years, so the three strata were not cleanly separated (Fig. 3(a)). Observed two-year risk (0.214) exceeded mean predicted risk (0.089) in the bridged calibration analysis, an observed-to-expected ratio of 2.40. Table II summarises two-year discrimination and calibration; the diagnostic calibration slope was 0.347 (95% CI, *−* 0.020 to 0.965) and was imprecisely estimated.

**Fig. 3.**
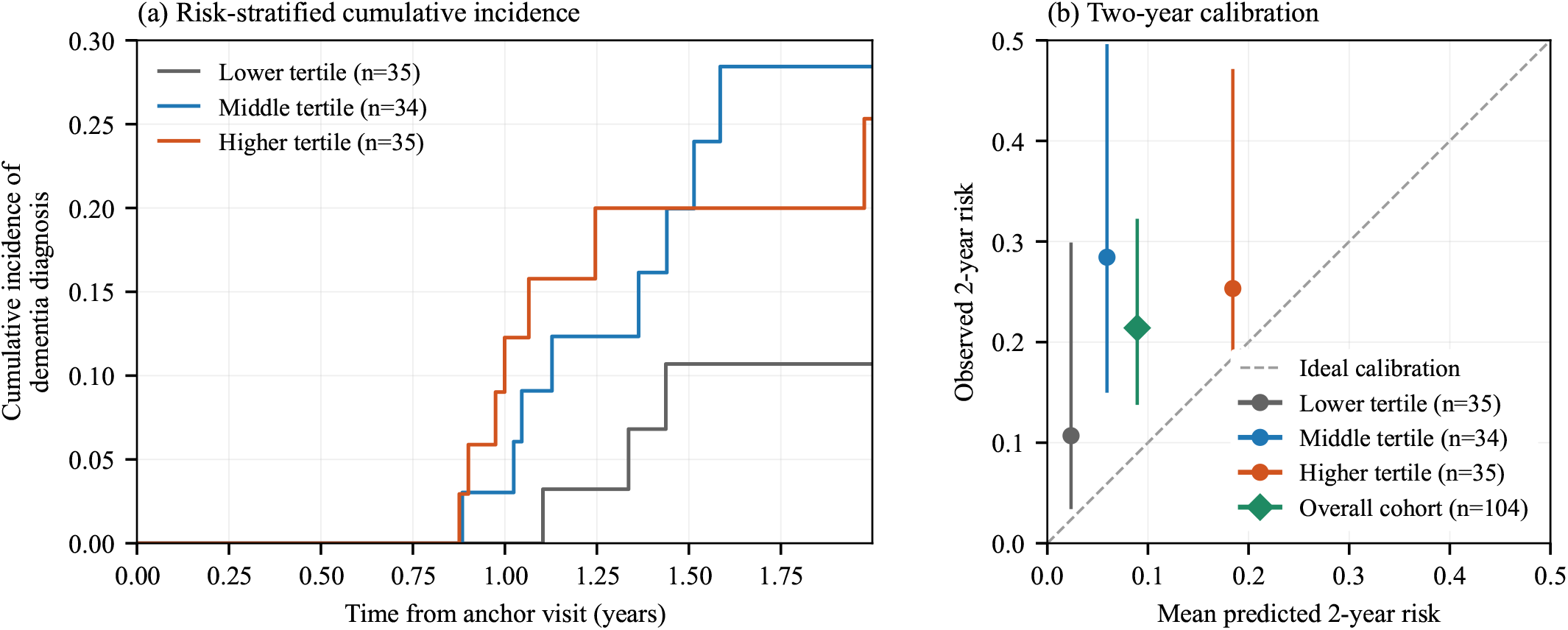
External validation in NACC of the frozen age-plus-p-tau217 model. (a) Cumulative incidence of a recorded dementia diagnosis by tertile of risk from the frozen bridged model; the middle and higher tertiles overlap and cross. (b) Two-year calibration comparing mean predicted risk with Kaplan–Meier observed risk; vertical bars denote 95% confidence intervals, the diagonal denotes ideal calibration, and the diamond denotes the overall cohort. Predictions were generated without model refitting or recalibration.

**TABLE II.** External validation of the frozen age-plus-p-tau217 model in NACC (*n*=104; 25 events)

|  | Bridged<br>analysis | Direct-scale<br>sensitivity analysis |
| --- | --- | --- |
| C-index (95% CI)<sup>a</sup> | 0.652<br>(0.526–0.765) | 0.654<br>(0.533–0.777) |
| Two-year AUC (95% CI) | 0.647<br>(0.498–0.791) | 0.648<br>(0.501–0.791) |
| Two-year Brier (95% CI) | 0.182<br>(0.113–0.255) | 0.173<br>(0.120–0.229) |
| Mean predicted two-year risk | 0.089 | 0.209 |
| Observed two-year risk (95% CI) | 0.214 (0.139–0.321) |  |
| Observed/expected (95% CI) | 2.40<br>(1.45–3.44) | 1.02<br>(0.63–1.46) |

<sup>a</sup>Includes assay-bridge uncertainty. Observed risk is a cohort property, common to both columns. Observed/expected is Kaplan–Meier observed risk divided by mean predicted risk.

Two-year performance was estimable, with 44 participants at risk and 18 events by that horizon; four-year performance was not, because no participants remained under observation at 1,461 days, the longest follow-up being 1,229 days.

Treating the NACC measurements as already equivalent to Fujirebio concentrations left discrimination essentially unchanged (C-index, 0.654) but changed absolute-risk calibration, raising mean predicted two-year risk to 0.209 and giving an observed-to-expected ratio of 1.02.

The secondary APOE analysis included 103 participants with complete APOE *ε*4 counts. The excluded participant was censored without a recorded dementia diagnosis, so all 25 incident dementia diagnoses were retained. In these participants, adding APOE *ε*4 allele count did not materially improve predictive performance. The paired difference in Harrell’s C-index was 0.000 (95% CI, *−* 0.009 to 0.009), and the differences in two-year AUC and two-year Brier score were 0.003 (95% CI, *−* 0.008 to 0.017) and 0.0003 (95% CI, *−* 0.0012 to 0.0005), respectively. Calibration was essentially unchanged.

In the post hoc age-at-index sensitivity analysis, replacing study-entry age with index-date age produced an out-of-fold C-index difference of *−* 0.002 in ADNI (95% CI, *−* 0.006 to 0.003). In NACC, the bridged C-index was 0.642, a paired difference of *−* 0.010 from the primary model (95% CI, *−* 0.027 to 0.008), and the observed-to-expected ratio was 2.35 compared with 2.40 using study-entry age.

## IV. Discussion

In this development cohort, the natural logarithm of plasma p-tau217 added substantially to age in ranking who reached a subsequent dementia diagnosis sooner, and the improvement remained large against a stronger demographic baseline that also contained sex and education. APOE *ε*4 count, sex and education added no confirmed incremental discrimination to the two-predictor model, while expansion with GFAP, NfL and A*β*42/A*β*40 likewise produced no material gain in the separate p-tau217–containing clinical benchmark. Together, these findings supported retention of the two-predictor model for external validation. AIC preferred the simpler model, and the advantage over the age-only baseline held across every supporting analysis.

Transported to NACC without refitting, the same two-predictor model showed moderate discrimination (0.652, against an out-of-fold 0.775 in development), but absolute risk did not transport as well: mean predicted two-year risk was less than half the observed Kaplan–Meier risk. Discrimination was similar in the bridged and direct-scale analyses, whereas absolute-risk calibration was sensitive to the measurement scale, as expected when rescaling a single predictor changes absolute risk more than rank ordering. The absence of meaningful incremental improvement after adding APOE *ε*4 allele count was consistent with the development results and supported retaining the age and p-tau217 model as the primary model.

The calibration slope remained low in both measurement-scale analyses, although its confidence interval was wide. Case mix, shifts in the predictor distribution, measurement error and scaling, the limited number of events and transportability of the coefficients could each contribute, and the external outcomes do not distinguish among them. These results describe rank ordering and short-term calibration; they do not establish clinical utility, a validated decision threshold, or a causal role for p-tau217, and estimate time to a recorded diagnosis rather than biological onset.

Several limitations bound this interpretation. The external cohort included only 25 events, resulting in wide confidence intervals for the external performance estimates; follow-up did not reach four years, so longer-horizon calibration could not be evaluated; and external validation required cross-platform harmonisation, so absolute-risk calibration was sensitive to the measurement scale. In development, final-model selection occurred within ADNI and was not repeated within each resampling split, so the internal estimates may not fully incorporate optimism from that selection. The primary model used age at study entry rather than at the plasma index date; replacing it with index-date age in a post hoc sensitivity analysis did not materially change performance, although the ADNI index age was reconstructed rather than directly recorded. The endpoint is dementia of any cause, etiologic diagnosis being recorded for too few events, and the reason for follow-up ending was not available, precluding assessment of independent censoring. Death was censored rather than modelled as a competing event; three NACC participants died during follow-up, so the Kaplan–Meier observed two-year risk of 0.214 may overstate real-world cumulative incidence, and the observed-to-expected ratio inherits that. Diagnoses are recorded at scheduled visits rather than continuously, so event times are interval-censored on each cohort’s visit schedule; NACC visits are approximately annual, visible in Fig. 3(a) as the absence of events before roughly 0.9 years, and the two-year risks are therefore comparable across cohorts only up to differences in visit timing. Because NACC specimen dates were unavailable, the linked clinical visit defined the index date; any lag between specimen collection and that visit could have affected calibration. The small external cohort precluded meaningful assessment of performance across demographic or clinical subgroups. The additional-biomarker comparison was a supporting analysis with only 85 events and required fold-specific handling of missing biomarker measurements, so the absence of improved discrimination should not be interpreted as evidence that these biomarkers have no biological or prognostic relevance.

## V. Conclusion

An age and plasma p-tau217 survival model developed in ADNI showed moderate discrimination in NACC (C-index, 0.652), but its 95% confidence interval (0.526–0.765) included values near chance; mean two-year risk was underestimated in the bridged primary analysis. Neither the supporting demographic, genetic, and biomarker extensions in ADNI nor the secondary APOE *ε*4 comparison in NACC materially improved discrimination, supporting retention of the two-predictor specification. Further validation in larger external cohorts with longer follow-up is needed to assess calibration and longer-term transport.

## Data Availability

ADNI data are available from the Alzheimer’s Disease Neuroimaging Initiative (adni.loni.usc.edu) and NACC Uniform Data Set data from the National Alzheimer’s Coordinating Center (naccdata.org), both to qualified investigators on application to the respective data-access committees. The harmonised plasma biomarker phenotypes are available from NIAGADS under accession NG00067.

## Ethics Statement

This study was a secondary analysis of previously collected, de-identified human participant data. Data from the Alzheimer’s Disease Neuroimaging Initiative (ADNI) were collected under protocols reviewed and approved by the institutional review boards or ethics committees of the participating institutions, and written informed consent was obtained from participants or their legally authorized representatives. National Alzheimer’s Coordinating Center (NACC) Uniform Data Set data were contributed by participating NIA-funded Alzheimer’s Disease Research Centers under their respective institutional protocols and consent procedures. The harmonized plasma biomarker phenotypes used for the external validation were obtained from the ADSP Phenotype Harmonization Consortium release NG00067.v20 and were analyzed only after linkage to the corresponding NACC clinical data.

The present study involved no recruitment, participant contact, intervention, or new collection or assay of biospecimens. Only de-identified data obtained through the applicable ADNI, NACC, and ADSP-PHC data-access procedures were analyzed. Data use was conducted in accordance with the applicable data-use agreements and restrictions, including requirements governing confidentiality and participant re-identification.

## Competing Interests

The authors declare no competing interests.

## Funding

The authors received no specific funding for the research, authorship, or publication of this study. Funding supporting the collection, maintenance, and harmonization of the ADNI, NACC, and ADSP-PHC data resources is described in the Acknowledgment and did not constitute direct funding of the present analysis.

## Acknowledgment

Data collection and sharing for the Alzheimer’s Disease Neuroimaging Initiative (ADNI) is funded by the National Institute on Aging (National Institutes of Health Grant U19 AG024904). The grantee organization is the Northern California Institute for Research and Education. In the past, ADNI has also received funding from the National Institute of Biomedical Imaging and Bioengineering, the Canadian Institutes of Health Research, and private sector contributions through the Foundation for the National Institutes of Health (FNIH) including generous contributions from the following: AbbVie, Alzheimer’s Association; Alzheimer’s Drug Discovery Foundation; Araclon Biotech; BioClinica, Inc.; Biogen; Bristol-Myers Squibb Company; CereSpir, Inc.; Cogstate; Eisai Inc.; Elan Pharmaceuticals, Inc.; Eli Lilly and Company; EuroImmun; F. Hoffmann-La Roche Ltd and its affiliated company Genentech, Inc.; Fujirebio; GE Healthcare; IXICO Ltd.; Janssen Alzheimer Immunotherapy Research & Development, LLC.; Johnson & Johnson Pharmaceutical Research & Development LLC.; Lumosity; Lundbeck; Merck & Co., Inc.; Meso Scale Diagnostics, LLC.; NeuroRx Research; Neurotrack Technologies; Novartis Pharmaceuticals Corporation; Pfizer Inc.; Piramal Imaging; Servier; Takeda Pharmaceutical Company; and Transition Therapeutics.

The NACC database is funded by NIA/NIH Grant U24 AG072122. NACC data are contributed by the NIA-funded ADRCs: P30 AG062429 (PI James Brewer, MD, PhD), P30 AG066468 (PI Oscar Lopez, MD), P30 AG062421 (PI Teresa Gomez-Isla, MD), P30 AG066509 (PI Thomas Grabowski, MD), P30 AG066514 (PI Mary Sano, PhD), P30 AG066530 (PI Helena Chui, MD, Arthur Toga, PhD), P30 AG066507 (PI Marilyn Albert, PhD), P30 AG066444 (PI David Holtzman, MD), P30 AG066518 (PIs Lisa Silbert, MD, Kevin Duff, PhD), P30 AG066512 (PI Thomas Wisniewski, MD), P30 AG066462 (PI Scott Small, MD), P30 AG072979 (PI David Wolk, MD), P30 AG072972 (PIs Charles DeCarli, MD, Rachel Whitmer, PhD), P30 AG072976 (PI Andrew Saykin, PsyD), P30 AG072975 (PI Julie Schneider, MD, MS), P30 AG072978 (PI Ann McKee, MD), P30 AG072977 (PI Robert Vassar, PhD), P30 AG066519 (PI Joshua Grill, PhD), P30 AG062677 (PIs Brad Boeve, MD, Ronald Petersen, MD, PhD), P30 AG079280 (PI Jessica Langbaum, PhD), P30 AG062422 (PI Gil Rabinovici, MD), P30 AG066511 (PI Allan Levey, MD, PhD), P30 AG072946 (PI Linda Van Eldik, PhD), P30 AG062715 (PI Sanjay Asthana, MD, FRCP), P30 AG072973 (PI Russell Swerdlow, MD), P30 AG066506 (PIs Glenn Smith, PhD, ABPP, David Lowenstein, PhD, Ranjan Duara, MD), P30 AG066508 (PIs Stephen Strittmatter, MD, PhD, Christopher Van Dyck, MD), P30 AG066515 (PI Victor Henderson, MD, MS), P30 AG072947 (PI Suzanne Craft, PhD), P30 AG072931 (PI Henry Paulson, MD, PhD), P30 AG066546 (PIs Sudha Seshadri, MD, Gladys Maestre, MD, PhD), P30 AG086401 (PI Erik Roberson, MD, PhD), P30 AG086404 (PI Gary Rosenberg, MD), P30 AG086403 (PI Angela Jefferson, PhD), P30 AG072958 (PIs Heather Whitson, MD, Gwenn Garden, MD, PhD), P30 AG072959 (PI Jagan Pillai, MD, PhD), P30 AG092752 (Ihab Hajjar, MD, MS).

The ADSP Phenotype Harmonization Consortium (ADSP-PHC) is funded by NIA (U24 AG074855, U01 AG068057 and R01 AG059716). The p-tau217 biomarker data used for external validation were from the February 2026 ADSP-PHC harmonized data release (ng00067.v20), accessed through the NACC Quick Access File data request system.

We thank Christopher T. Lee, Ph.D., for faculty oversight and support of the NACC data-access process, and Ifunanya Okoroma for guidance throughout the project.

## Notes

### Competing Interest Statement

The authors have declared no competing interest.

### Author Declarations

IRB of University of California, San Diego waived ethical approval for this work

## References

[1] J. Gao, L. Liu, Z. Yang, and J. Fan, “Predictors of transition from mild cognitive impairment to normal cognition and dementia,” Behavioral Sciences, vol. 15, no. 11, art. 1552, 2025, doi: 10.3390/bs15111552.

[2] B. Arslan, H. Zetterberg, and N. J. Ashton, “Blood-based biomarkers in Alzheimer’s disease – moving towards a new era of diagnostics,” Clinical Chemistry and Laboratory Medicine, vol. 62, no. 6, pp. 1063–1069, 2024, doi: 10.1515/cclm-2023-1434.

[3] R. Ossenkoppele, R. van der Kant, and O. Hansson, “Tau biomarkers in Alzheimer’s disease: towards implementation in clinical practice and trials,” The Lancet Neurology, vol. 21, no. 8, pp. 726–734, 2022, doi: 10.1016/S1474-4422(22)00168-5.

[4] S. Palmqvist et al., “Discriminative accuracy of plasma phospho-tau217 for Alzheimer disease vs other neurodegenerative disorders,” JAMA, vol. 324, no. 8, pp. 772–781, 2020, doi: 10.1001/jama.2020.12134.

[5] E. H. Thijssen et al., “Plasma phosphorylated tau 217 and phosphorylated tau 181 as biomarkers in Alzheimer’s disease and frontotemporal lobar degeneration: a retrospective diagnostic performance study,” The Lancet Neurology, vol. 20, no. 9, pp. 739–752, 2021, doi: 10.1016/S1474-4422(21)00214-3.

[6] S. Lehmann et al., on behalf of the BALTAZAR study group, “Clinical value of plasma ALZpath pTau217 immunoassay for assessing mild cognitive impairment,” Journal of Neurology, Neurosurgery & Psychiatry, vol. 95, no. 11, pp. 1046–1053, 2024, doi: 10.1136/jnnp-2024-333467.

[7] R. F. Buckley et al., “Prognostic value of blood-based p-tau217 levels for progression to cognitive impairment,” JAMA, published online July 14, 2026, doi: 10.1001/jama.2026.12556.

[8] N. C. Cullen et al., “Individualized prognosis of cognitive decline and dementia in mild cognitive impairment based on plasma biomarker combinations,” Nature Aging, vol. 1, pp. 114–123, 2021, doi: 10.1038/s43587-020-00003-5.

[9] R. C. Petersen et al., “Alzheimer’s Disease Neuroimaging Initiative (ADNI): clinical characterization,” Neurology, vol. 74, no. 3, pp. 201–209, 2010, doi: 10.1212/WNL.0b013e3181cb3e25.

[10] G. S. Collins, J. B. Reitsma, D. G. Altman, and K. G. M. Moons, “Transparent reporting of a multivariable prediction model for individual prognosis or diagnosis (TRIPOD): the TRIPOD statement,” Annals of Internal Medicine, vol. 162, no. 1, pp. 55–63, 2015, doi: 10.7326/M14-0697.

[11] D. R. Cox, “Regression models and life-tables,” Journal of the Royal Statistical Society: Series B (Methodological), vol. 34, no. 2, pp. 187–202, 1972, doi: 10.1111/j.2517-6161.1972.tb00899.x.

[12] F. E. Harrell Jr., K. L. Lee, and D. B. Mark, “Multivariable prognostic models: issues in developing models, evaluating assumptions and adequacy, and measuring and reducing errors,” Statistics in Medicine, vol. 15, no. 4, pp. 361–387, 1996, doi: 10.1002/(SICI)1097-0258(19960229)15:4<361::AID-SIM168>3.0.CO;2-4.

[13] P. M. Grambsch and T. M. Therneau, “Proportional hazards tests and diagnostics based on weighted residuals,” Biometrika, vol. 81, no. 3, pp. 515–526, 1994, doi: 10.1093/biomet/81.3.515.

[14] P. K. Andersen and M. Pohar Perme, “Pseudo-observations in survival analysis,” Statistical Methods in Medical Research, vol. 19, no. 1, pp. 71–99, 2010, doi: 10.1177/0962280209105020.

[15] P. C. Austin, F. E. Harrell Jr., and D. van Klaveren, “Graphical calibration curves and the integrated calibration index (ICI) for survival models,” Statistics in Medicine, vol. 39, no. 21, pp. 2714–2742, 2020, doi: 10.1002/sim.8570.

